# Circulation of Fluconazole-Resistant *C. albicans, C. auris* and *C. parapsilosis* Bloodstream Isolates Carrying Y132F, K143R or T220L Erg11p Substitutions in Colombia

**DOI:** 10.1101/2022.11.30.22282939

**Authors:** Andres Ceballos-Garzon, Ana Peñuela, Sandra Valderrama-Beltrán, Yerly Vargas-Casanova, Beatriz Ariza, Claudia M. Parra-Giraldo

## Abstract

Although *Candida* spp., is a common cause of bloodstream infections and is often associated with high mortality rates, its resistance to antifungal drugs, and the molecular mechanisms involved have been poorly studied in Colombia. Here, 123 bloodstream isolates of *Candida* spp. were collected. MALDI-TOF MS identification and fluconazole (FLC) susceptibility patterns were assessed on all isolates. Subsequently, sequencing of *ERG11, TAC1* or *MRR1*, and efflux pumps were performed for resistant isolates. Out of 123 clinical strains, *C. albicans* accounted for 37.4%, followed by *C. tropicalis* 26.8%, *C. parapsilosis* 19.5%, *C. auris* 8.1%, *C. glabrata* 4.1%, *C. krusei* 2.4% and *C. lusitaniae* 1.6%. Resistance to FLC reached 18%. Erg11 amino acid substitutions associated with FLC-resistance (Y132F, K143R or T220L) were found in 58% of 19 FLC-resistant isolates. Furthermore, novel mutations were found in all genes studied. Regarding efflux pumps, 42% of 19 FLC-resistant *Candida* spp strains showed significant efflux activity. Finally, six of the 19 FLC-resistant isolates neither harbored resistance-associated mutations nor showed efflux pump activity. Although *C. albicans* remain the most predominant species, non-*C. albicans* species comprise a high proportion (62.6%). Among FLC-resistant species, *C. auris* (70%) and *C. parapsilosis* (25%) displayed the highest percentages of resistance. In 68% of FLC-resistant isolates, a mechanism that could explain their phenotype was found (e.g. mutations, flux pump activity or both). We provide evidence that endemic isolates harbor amino acid substitutions related with resistance to one of the most used molecules in the hospital setting, with Y132F being the most frequently detected one.

## Introduction

Invasive fungal infections (IFIs) due to *Candida* species are a frequent and life-threatening condition in hospital settings worldwide, and are often associated with high morbidity and mortality [1]. *Candida albicans* is the most isolated species, with close to a 30% mortality rate in candidemia [2]. However, non-*albicans Candida* species (NACS) such as *C. glabrata* and *C. parapsilosis* have emerged as a common cause, becoming the second or third most frequent species depending on geography, patient underlying condition, and age. For these species, the associated mortality rate is about 50% and 28%, respectively [3,4]. Another important NACS causing candidemia is *C. auris*, whose mortality rate ranges from 40% to 60% according to some studies, despite the fact its prevalence is unclear [5].

While current therapeutic options for IFIs are limited to only three classes of drugs (i.e., polyenes, azoles and echinocandins), the emergence of resistant strains to some of these molecules is even more concerning. For decades, azoles have been the most frequently used antifungal for treating *Candida* infections [6,7]. Although treatment with azoles can be effective, long-term use of fluconazole (FLC) has led to the emergence of *Candida* strains with decreased susceptibility. In the case of *Candida* spp, the molecular mechanisms behind FLC-resistance have been relatively well characterized [8,9].

Unlike *C. auris* which is often resistant to FLC, *C. albicans* and *C. parapsilosis* isolates were thought to be universally susceptible to FLC, but recent studies show an increased resistance. For example, a multicenter laboratory-based survey of candidemia conducted in South Africa indicates that more than half of *C. parapsilosis* isolates 62% (332/531) are resistant to FLC [10]. In addition, studies in Brazil, India, Kuwait, South Korea, Spain, Turkey, and the United States, as well as a recent global study, confirmed the emergence of FLC-resistance in *C. parapsilosis* [11–15]. Regarding *C. albicans*, this species exhibits lower levels of azole resistance. However, resistant isolates have been reported from many countries around the world, including Colombia [16–18].

The main FLC-resistance mechanisms are associated with *i*) up-regulation of drug transporters, *ii*) alteration or up-regulation of the gene encoding the enzyme being targeted, which decreases binding affinity for the drug and increases concentration of the enzyme target, *iii*) alterations in the ergosterol synthetic pathway and *iv*) activation of pathways involved in the stress response, such as the Ras/cAMP/PKA pathway, calmodulin/calcineurin pathway (CaM/CaL), and mitogen-activated protein kinase (MAPK) signaling pathways [19,20]. However, the most studied ones are described below.

The ATP-binding cassette (ABC) and the major facilitator superfamily (MFS) transporters are responsible for lowering the accumulation of azoles inside the yeast cell by translocating compounds actively across the cell membrane [21]. Overexpression of genes encoding drug transporters, e.g., Cdr1/2-ABC and Mdr1/Flu1-MFS, among resistant isolates of *Candida* species is predominantly due to gain-of-function (GOF) mutations in genes encoding zinc cluster transcription factors, such as *TAC*1 (transcriptional <u>a</u>ctivator of <u>C</u>DR genes) and *MRR*1 (<u>m</u>ultidrug resistance <u>r</u>egulator). For instance, GOFs in *TAC*1 (T225A, R693K, A736V, H741, N972D, G980E, N997D) and *MRR*1 (I283R, R479K, G583R, V854A, K873N), lead to overexpression of *CDR*1 and *MDR*1 in *C. albicans and C. parapsilosis*, respectively [22,23].

Overexpression of *ERG*11, the gene encoding lanosterol 14α-demethylase, the azole target, contributes directly to resistance as the increased abundance of the target requires higher drug doses for inhibition. Activating mutations in the gene encoding the transcription factor Upc2, which up-regulates most ergosterol biosynthesis genes, and the formation of an isochromosome with two copies of the left arm of chromosome 5 [i(5L)], or by duplication of the whole chromosome, on which *ERG*11 resides, are responsible for *ERG*11 overexpression [24]. Furthermore, point mutations in the *ERG*11 alter the 3D conformation of Erg11 and reduce its affinity for FLC. Some of the most frequent amino acid substitutions reported are Y132F and K143R substitutions, described in *C. albicans, C. parapsilosis* and *C. auris* [15,25,26].

Although *Candida* isolates from individual institutions may not be representative of the data of a country, such studies can provide a useful baseline snapshot of species distribution and antifungal susceptibility for candidemia in resource-limited settings [10]. In Colombia, there is a lack of data available about antifungal resistance and its molecular mechanisms in *Candida* spp. Therefore, this study aimed to investigate the prevalence of resistance and to describe the mechanisms behind FLC-resistance in a collection of bloodstream isolates.

## Materials and methods

### Study design

The study was a single-center retrospective analysis. One hundred twenty-three bloodstream isolates of *Candida* spp. obtained from hospitalized patients (2016-2020) of the San Ignacio Hospital in Bogota, Colombia were included. Prior to storage at −80 °C, yeast from blood cultures submitted for routine work-up to the Clinical Microbiology Laboratory were primarily identified using MicroScan (MicroScan WalkAway-96 Plus, Siemens, Deerfield, IL, USA) or VITEK 2 system (bioMérieux, Marcy-l’Etoile, France), and further characterized (this study) using the MALDI-TOF Biotyper system (Bruker Daltonik, Bremen, Germany).

### 2.2 MALDI-TOF MS

Isolates were streaked from a glycerol stock onto Sabouraud dextrose agar (SDA) and grown for 24–36 h at 35 °C. Protein extraction was performed using formic acid/ethanol method, according to the Bruker Daltonics’ protocol. The protein mass spectra were analysed using the Flex Control software and the MALDI Biotyper version 3.1 7311 reference spectra (main spectra) (Bruker Daltonics, Bremen, Germany). MALDI-TOF MS results were obtained according to the manufacturer’s technical specifications, as follows: correct genus and species identification (≥2.0), correct genus identification (1.7–2.0), and no reliable identification (< 1.7). All clinical isolates had a score above 2.0 [27].

### 2.3 Antifungal susceptibility testing

Susceptibility to FLC (Sigma-Aldrich, St. Louis, MO, USA) was conducted using the Clinical and Laboratory Standards Institute broth microdilution method (CLSI-BMD), following the M27-A3 document [28]. Briefly, isolates were suspended in 1 mL phosphate buffered saline (PBS) and diluted in liquid RPMI 1640 medium to 10^3^ cells/mL in a 96-well plate, containing a gradient of two-fold dilutions per step of antifungal, with the first well containing no drug. Minimum inhibitory concentrations (MICs) were visually, and densitometry determined as the lowest concentration of drug that caused a significant diminution (MIC/2 or >50%) compared with that of the drug-free growth control after 24 h of incubation. Quality control was ensured by testing the CLSI-recommended strains *C. parapsilosis* ATCC 22019 and *C. krusei* ATCC 6258. For NACS isolates the CLSI breakpoints were applied (resistance to FLC was set at *C. albicans, C. tropicalis, and C. parapsilosis ≥*8μg/mL and *C. glabrata ≥*64μg/mL) [29]. In the case of *C. auris*, the FLC breakpoint recommended by the US Centers for Disease Control and Prevention (CDC) was used (≥32 μg/mL) [30]. The MIC data obtained under routine conditions for amphotericin B (AMB), caspofungin (CAS), itraconazole (ITC), and voriconazole (VRC) by Etest® (bioMérieux, Marcy-l’Étoile, France) and VITEK®2 (bioMérieux) are presented in **Table S1**.

### Sequencing analysis of Erg11, Tac1 and Mrr1-encoding genes

All isolates displaying resistance to FLC, and one susceptible isolate of each species were subjected to a single-tube PCR method to amplify and sequence the coding region of the *ERG11, TAC*1 or *MRR*1 genes (both strands) using the primers indicated in **Table S2**. The PCR products were purified and sequenced using a SeqStudio genetic analyzer capillary sequencer (Applied Biosystems). The sequencing results were analyzed by BLAST and compared with the published GenBank sequences: *C. albicans* AY856352.1 (*ERG*11), DQ393587 (*TAC*1), and *C. parapsilosis* GQ302972 (*ERG*11), HE605205 (*MRR*1). For *C. auris*, sequences download from the Candida genome database (candidagenome.org) were used, i.e., B9J08_001448 (*C. auris* B8441, *ERG*11) and B9J08_004820 (*TAC*1b). All sequences (FLC-susceptible and resistant clinical isolates plus reference strains) were aligned, and the dataset was used to construct a Neighbor-Joining phylogenetic tree using Maximum Composite Likelihood settings by using Molecular Evolutionary Genetics Analysis Version 11 (MEGA11) [31,32]. Codon positions included were 1st + 2nd + 3rd + Noncoding. All positions containing gaps and missing data were eliminated. Evaluation of branch support was performed by Bootstrap statistical analysis with 1000 replicates [31].

### Analysis of rhodamine 6G efflux

ABC transporter-mediated efflux was determined using rhodamine 6G (Sigma-Aldrich, USA) as previously described [33]. The fluorescence of the released R6G was measured at 530 nm, with an emission at 560 nm in an automated plate reader (Model 550 Microplate Reader Bio-Rad, Milan, Italy). Measurements were made before (basal) and after the addition of 20 mM glucose. Using a R6G calibration curve, the fluorescence intensity was converted into concentration.

### Statistics

Experiments were performed in triplicate, in three independent experimental sets. The results were analyzed statistically by the Analysis of Variance One-Way ANOVA. All the analyses were performed using GraphPad Prism version 9 software. In all analyses, p values of 0.05 or less were considered statistically significant.

## Results

### Identification and Antifungal susceptibility testing of clinical isolates

The *Candida* species distribution from the 123 blood samples was as follows: *C. albicans*, 46 (37.4%); *C. tropicalis*, 33 (26.8%); *C. parapsilosis*, 24 (19.5%); *C. auris*, 10 (8.1%); *C. glabrata*, 5 (4.1%); *C. krusei*, 3 (2.4%); and *C. lusitaniae*, 2 (1.6%). Although *C. albicans* was the most prevalent species, accounting for 37.4%, the NACS group comprised 62.6% of the isolates identified. Concerning susceptibility, when applying the CLSI and CDC breakpoints, 22 out of 123 (18%) isolates displayed *in vitro* resistance to FLC, among them, six of *C. albicans*, six of *C. parapsilosis* and seven of *C. auris* (19/23). Moreover, three isolates of *C. krusei -*as *C. krusei* is assumed to be intrinsically resistant to FLC-, were not included within the molecular study. In contrast, all isolates of *C. tropicalis, C. glabrata*, and *C. lusitaniae* were FLC-susceptible **Figure 1**.

**Figure 1.**
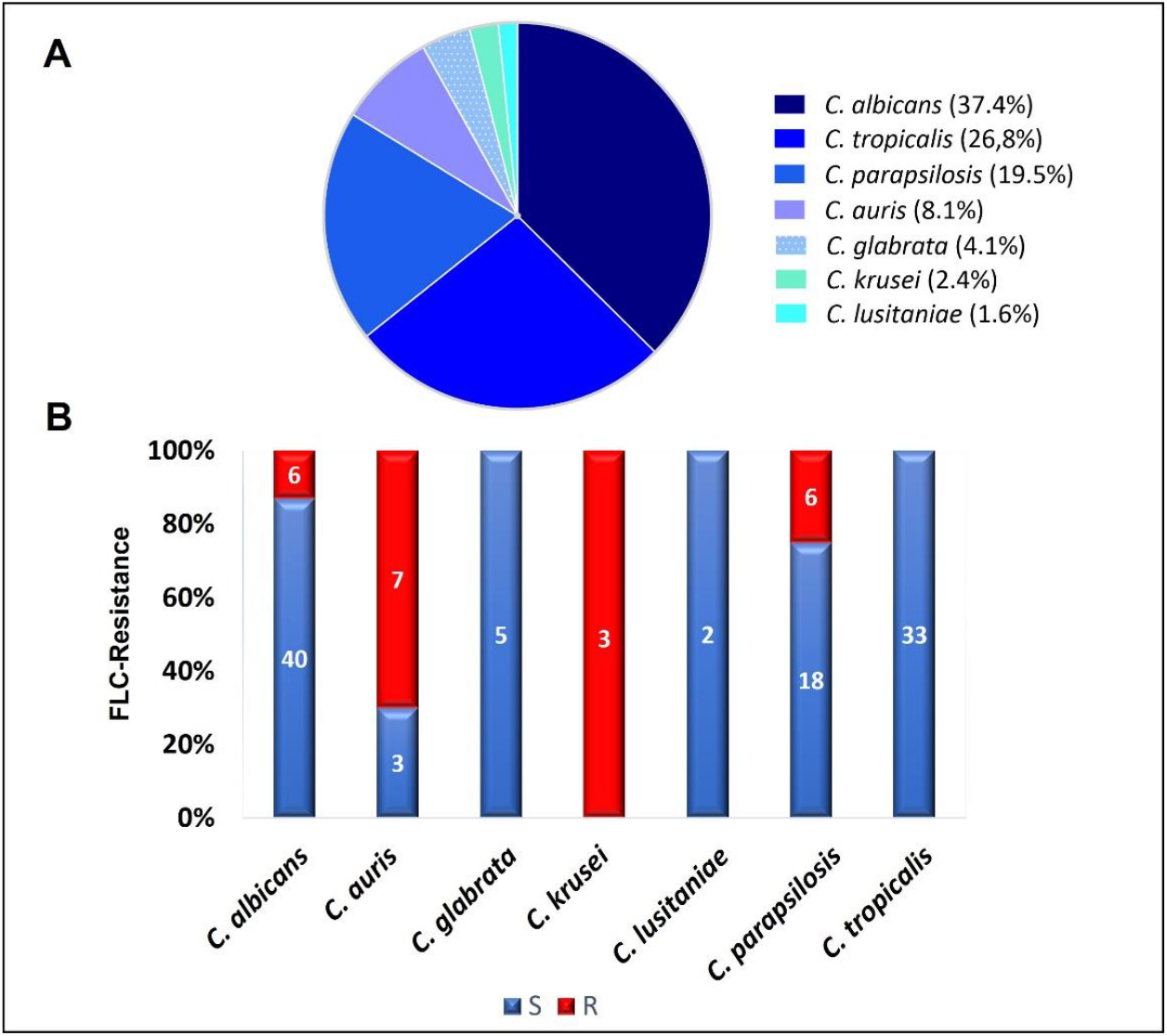
Species distribution and overall fluconazole susceptibility results. **A)** Distribution of the 123 species identified. **B**) Percentage of fluconazole resistance. The number of isolates is indicated inside the bars. Susceptible (S) isolates are depicted in blue and resistant (R) isolates in red.

The MICs values obtained for FLC are shown in **Table 1**. The MIC_90_ values (MICs at which ≥90% of strains are inhibited) for the four most frequent species found were: *C. albicans* 128μg/mL, *C. tropicalis* 2μg/mL, *C. parapsilosis* 32μg/mL and *C. auris* >128μg/mL. As expected, *C. auris* presented the highest MIC_90_ values. In addition, the range of FLC MICs was narrower for *C. auris* and *C. glabrata* than for the other species. On the other hand, *C. tropicalis* showed reduced susceptibility to FLC (MIC_90_= 2μg/mL). Regarding VRC MICs (data obtained by Etest), 10 out of 19 FLC-resistant isolates, excluding *C. krusei*, were VRC cross-resistant. The highest MICs were observed for *C albicans* (32 μg/mL) **Table S3**.

**Table 1.**
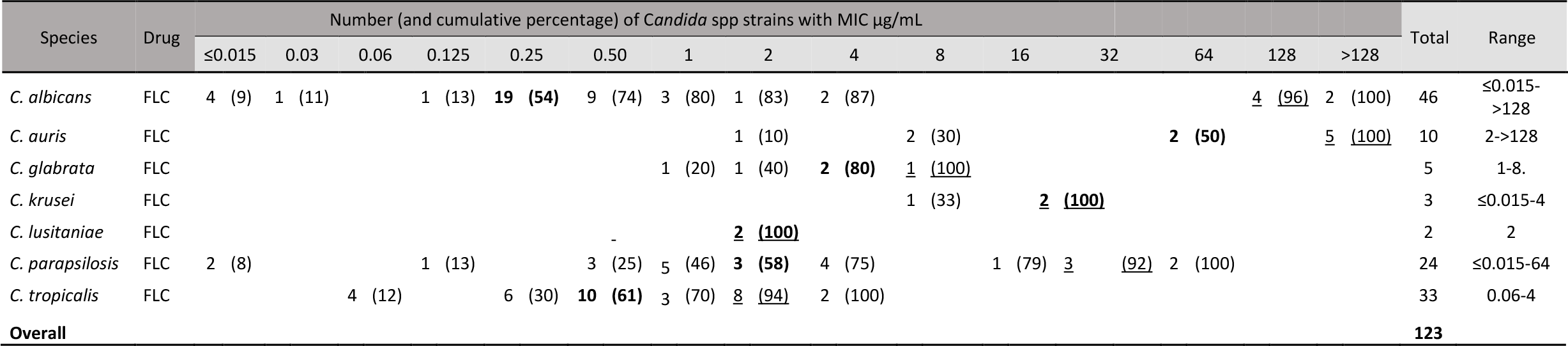
Antifungal activity of fluconazole drug against *Candida* spp (n = 123) performed by CLSI. MIC_50_ and MIC_90_ values (MICs at which ≥50% and ≥90% of the strains are inhibited, respectively) are depicted in bold and underlined, respectively.

### Detection of mutations in Erg11, Tac1 and Mrr1-encoding genes of 19 FLC-resistant isolates

By comparing the *ERG*11 coding region of *C. albicans* (CAAL, A-F), *C. auris* (CAAU, A-G) *and C. parapsilosis* (CAPA, A-F) FLC-resistant isolates with that of our FLC-susceptible and the published wild-type sequences, we identified 20, 16, and three mutations, respectively. As expected, some silent mutations that do not change the protein sequence were identified (data not shown). The remaining *ERG*11 mutations which resulted in amino acid changes are shown in **Figure 2, Table S4**. Among the nonsense mutations (*C. albicans* 6; *C. auris* 5; and *C. parapsilosis* 2), three amino acid substitutions related with FLC-resistance (T220L, Y132F, K143R) were found from which Y132F was the most detected. Additionally, five amino acid substitutions previously described in FLC-susceptible isolates were observed (D116E, K128T, K177R, N335S, E343D). To the best of our knowledge, four amino acid substitutions (K22E, Q38T, F72V, Q77S) have not been previously reported. Overall, eight of the 19 FLC-resistant isolates did not have resistance-associated substitutions.

**Figure 2.**
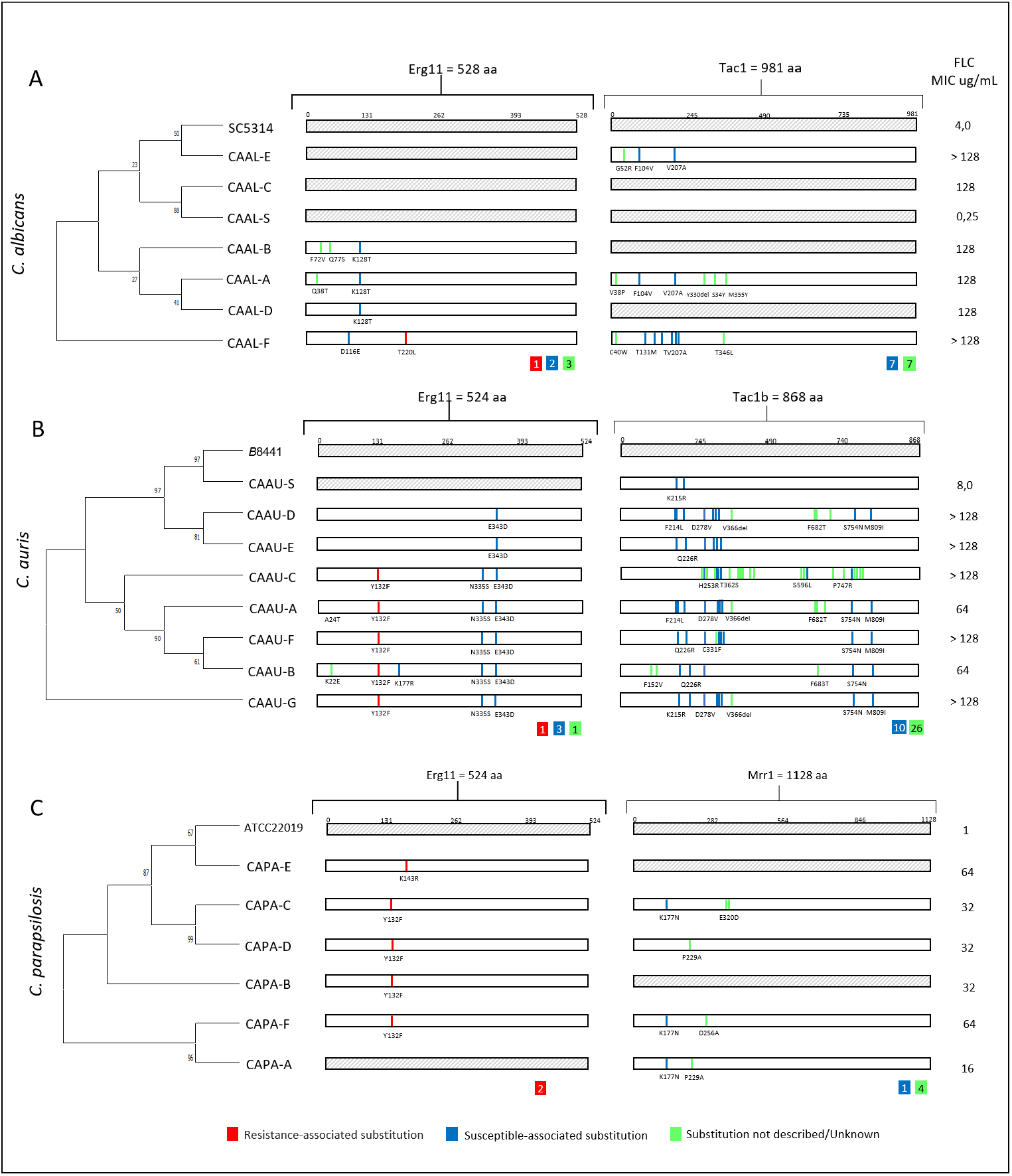
Phylogenetic tree and amino acid substitutions found in fluconazole-resistant isolates of **A**) *C. albicans* (CAAL), **B**) *C. auris* (CAAU) and **C**) *C. parapsilosis* (CAPA). aa., amino acids. FLC: fluconazole; MIC: minimal inhibitory concentration. CA-S., *Candida* FLC-susceptible. Inside the colored squares, the number of substitutions found is indicated.

On the other hand, except for isolate CAAL-A, which harbored a K128T substitution in only one *ERG*11 allele, other isolates were homozygous for mutations in the *ERG*11 allele. In the phylogenetic relationship among FLC-resistant, susceptible isolates and reference strains, a cluster of isolates carrying the substitutions Y132F in *C. auris* was observed, as well as in the FLC-resistant *C. albicans* isolates harboring K128T substitution. In addition, susceptible and resistant isolates without resistance-associated *ERG*11 mutations (i.e., T220L, Y132F) from these species were clustered **Figure 2A-B**.

Concerning *TAC1* (*C. albicans* and *C. auris*) and *MRR1* (*C. parapsilosis*) genes, no mutations previously associated with FLC-resistance were found. However, 17 Tac1 (*C. albicans*, n=7; *C. auris*, n=10) and Mrr1 (*C. parapsilosis*, n*=*1) amino acid substitutions previously described in FLC-susceptible isolates were found. Additionally, there were 37 unreported substitutions in Tac1 (*C. albicans*, n=7; *C. auris, n=*26) and Mrr1 (*C. parapsilosis, n=4*) **Figure 2, Table S3**.

### Efflux Pumps activity

To gain further insights into the mechanisms of azole resistance in the clinical isolates the activity of efflux pumps was evaluated using rhodamine 6G, which uses the same membrane ABC transporters (Cdr1p and Cdr2p) as FLC in *Candida*. Among the 19 FLC-resistant isolates, eight of them showed significant active efflux of rhodamine-6G after addition of glucose: two of them belonging to *C. albicans* (CAAL-C and CAAL-E); four to *C. auris* (CAAU-A, CAAU-B, CAAU-C and CAAU-G) and two to *C. parapsilosis* (CAPA-C, CAPA-F) **Figure 3, Figure 1S**. Interestingly, no amino acid substitutions associated with FLC-resistance were identified in both *C. albicans* isolates (CAAL-C, 128μg/mL), (CAAL-D, 128μg/mL). Regarding the remaining isolates that showed efflux pump activity, *C. auris* (CAAU-A, CAAU-B, CAAU-C, CAAU-G) and *C. parapsilosis* (CAPA-C, CAPA-F) harbored the Y132F substitution.

**Figure 3.**
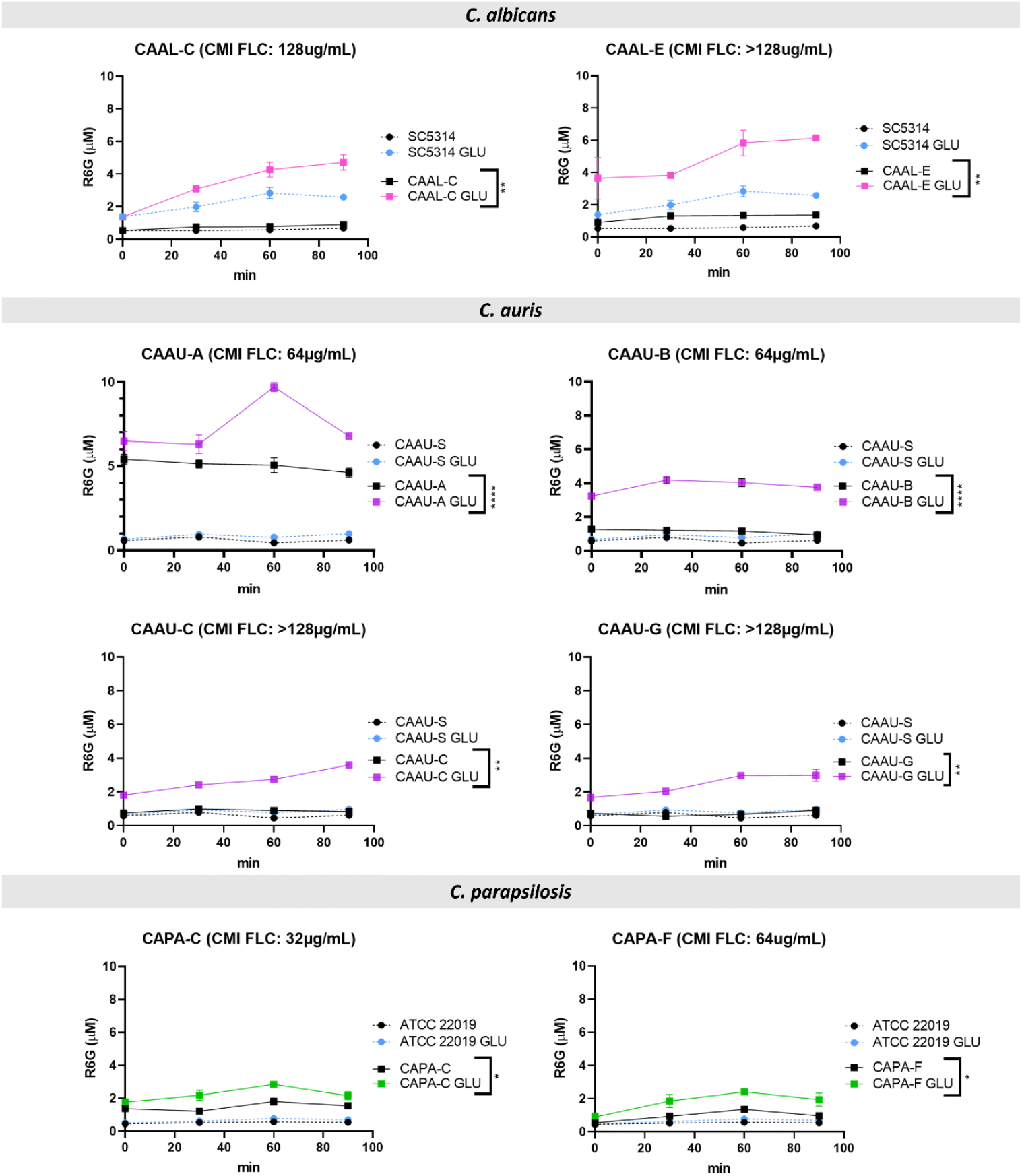
Rhodamine 6G (R6G) efflux over time among fluconazole-resistant isolates. Colored lines indicate the concentration of R6G released after the addition of 20 mM glucose (GLU). Data are means ± SD from three experiments. *p < 0.05, **p < 0.01, ***p < 0.001. ****p≤ 0.0001. FLC: fluconazole; MIC: minimal inhibitory concentration.

In summary, 11/19 resistant isolates harbored an amino acid substitution associated with FLC-resistance; 8/19 displayed efflux pumps activity; 6/19 had both amino acid substitutions and efflux pump activity; and 3/19 isolates did not exhibit any of the mechanisms that could explain their resistant phenotype **Figure 4**.

**Figure 4.**
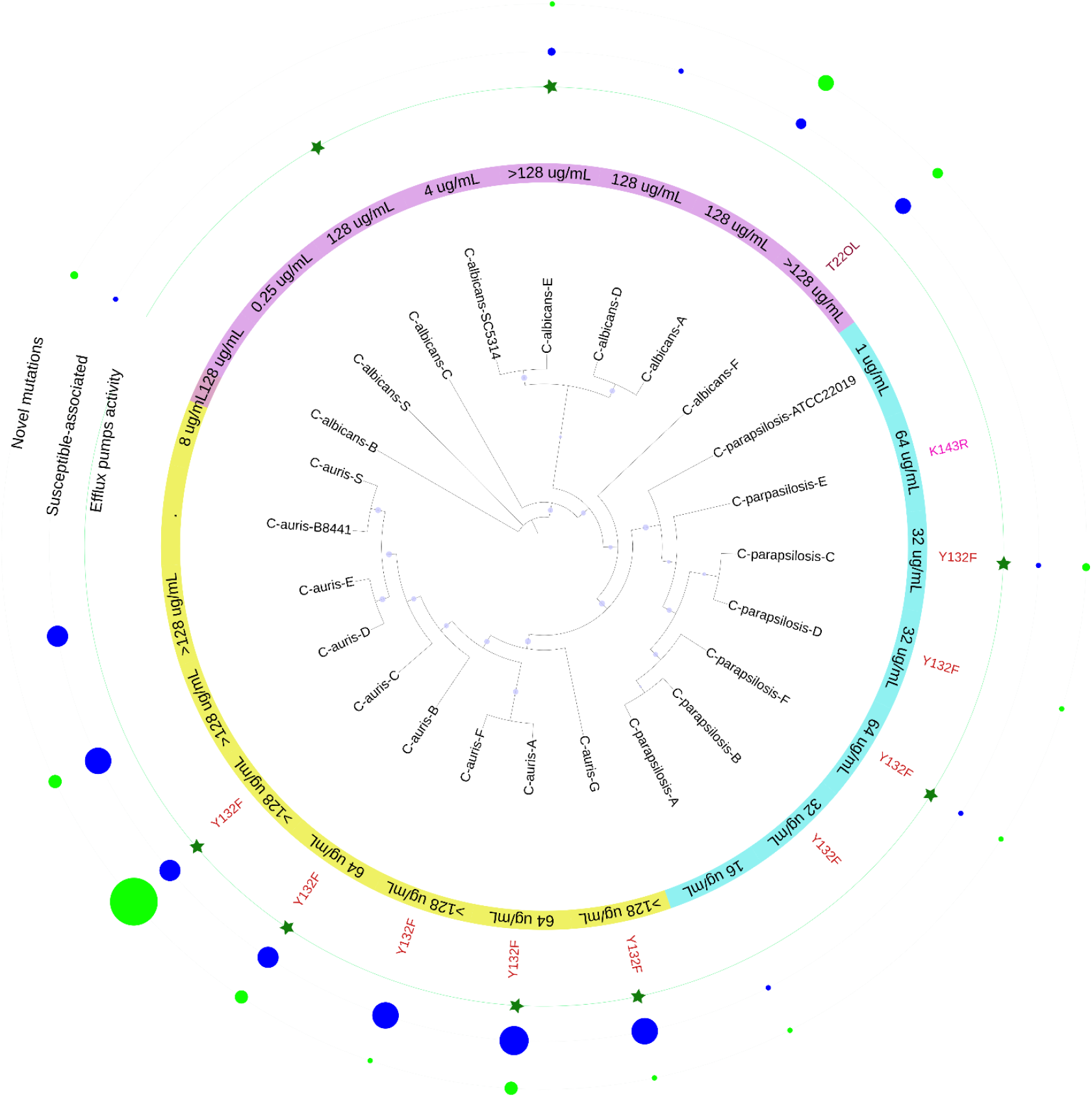
Schematic representation of the results obtained in this study for the 19 FLC-resistant isolates. In purple *C. albicans*, light blue *C. parapsilosis* and yellow *C. auris* isolates with their respective MICs against FLC. In red font, the mutations found. The green stars indicate isolates with significant efflux pump activity. The size of the circle reflects the number of mutations, those not associated with FLC resistance (blue) and the new mutations (light green).

## Discussion

Our study agrees that the three species most found in bloodstream infections in Latin America are *C. albicans, C. tropicalis* and *C. parapsilosis* [34,35]. However, the percentage of *C. tropicalis* (26.8% vs. 13.6%) and *C. parapsilosis* (19.5% vs. 13.6%) found differs from that observed in the first decade of the 2000s in Colombia [36]. Similar results were found for species distribution in studies conducted at the same hospital between 2003 and 2014, however, we observed a slight increase in NACS (62, 6% vs. 58%) and the presence of *C. auris*, which was not described in these studies [37,38]. In the region, a study that evaluated susceptibility of the species identified in Colombia, Ecuador, and Venezuela, found a percentage of resistance to FLC of 6.8% [39]. This shows that there is an important change in the distribution and rate of resistance, which in our study reached 18%. Nevertheless, the majority of *C. albicans* isolates were azole susceptible, thus the observed resistance percentage is mainly attributed to the presence of *C. auris*, which agrees with previous studies [35,40]. Furthermore, it is noteworthy that 25% of the *C. parapsilosis* isolates were resistant to FLC, which confirms the increase of resistance in this species **Figure 5** [41]. High azole resistance rates have been reported for this species in other single-center studies conducted in Brazil (67.9%), Italy (33%), France (9.2%), Mexico (54%), Saudi Arabia (33%), Spain (13.6%), South Africa (78%) or Turkey (26.4%) [11,13,42–46].

**Figure 5.**
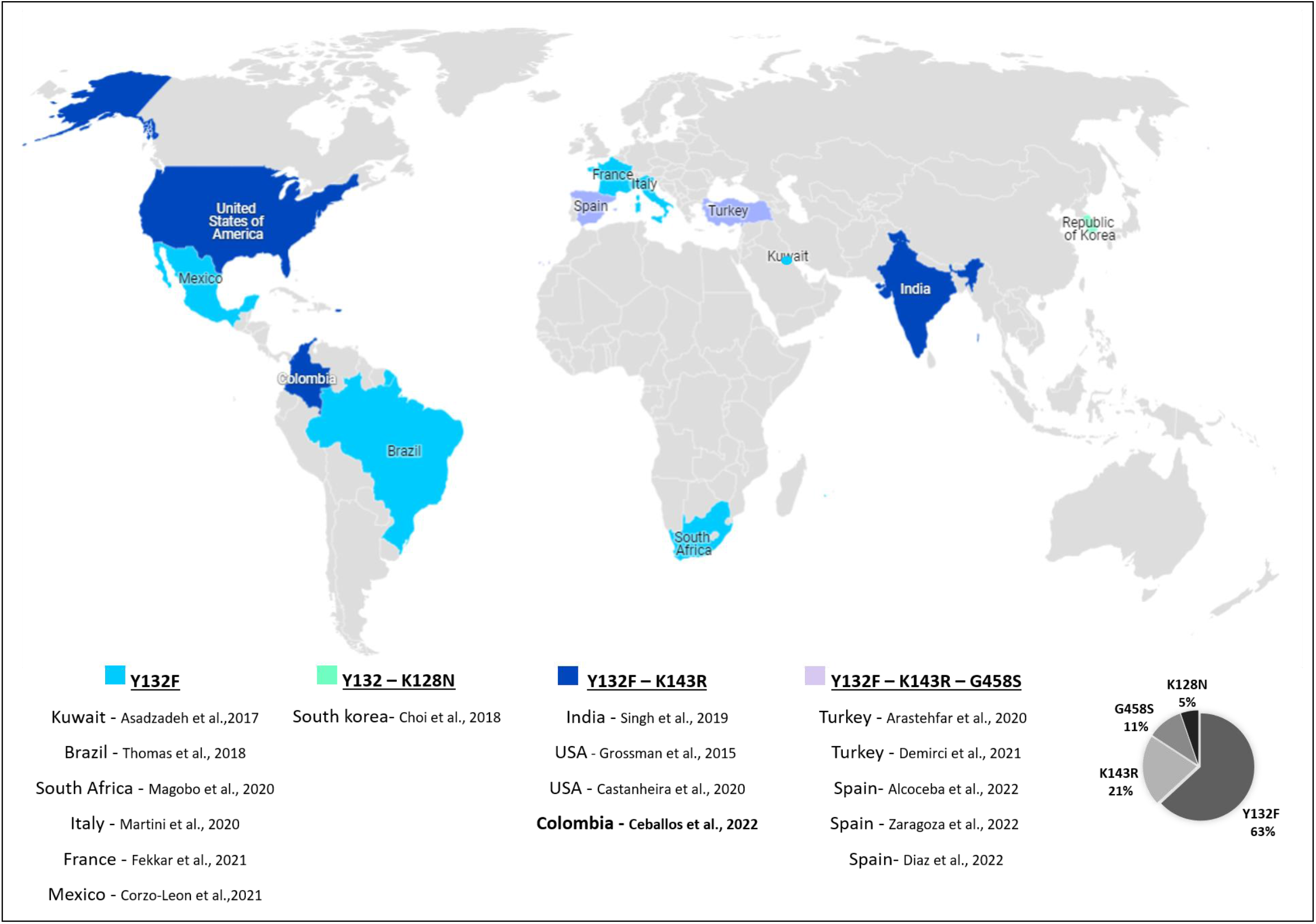
Countries reporting Erg11 substitutions in azole-resistant *C. parapsilosis*.

Overall, the global data indicate that *C. albicans* remains the predominant species identified in Candida infections. Herein, *C. albicans* was the most predominant species, nevertheless, NACS accounted for a high proportion (62.6%). Although reported resistance rates vary from study to study, the surveillance data collected suggest that azole resistance rates for *C. albicans* remain low [35,47]. Remarkably, in this study, the percentage of FLC resistance in *C. albicans* was 15%, which is relatively high.

However, *C. albicans* infections in the bloodstream pose a considerable threat in immunocompromised populations and the associated high mortality remains a major problem in the clinical setting. Therefore, *C. albicans* should not be overlooked as a serious public health threat.

The acquisition of azole resistance is a serious concern given the limited number of molecules available for the treatment of IFIs. Moreover, this is much more worrying in resource-limited regions where FLC is the only available therapy [48]. As previously described, azole resistance is mainly conferred by mutations in the *ERG*11 gene and by the activity of efflux pumps. The *ERG*11 gene is highly polymorphic and more than 140 amino acid substitutions have been reported, indicating that this protein is very permissive to conformational changes. Most substitutions occur in three amino acid hotspot regions (105-165, 266-287 and 405-488), although mutations outside these regions can also be found [49,50].

In the present study, 11 of the 19 FLC-resistant isolates harbored mutations which have been previously described in resistant isolates (T220L, Y132F, K143R) [11,51]. Notably, the T220L substitution was observed in one *C. albicans* isolate. In *C. auris*, five isolates carried the Y132F, and in the case of *C. parapsilosis* four isolates harbored the Y132F and one the K143R substitution.

The substitution of lysine for threonine at position 128 (K128T) was found in CAAL-A, CAAL-B and CAAL-D isolates, which is consistent with the findings of Cernicka *et al*., and Peron *et al*., who identified this amino acid substitution in FLC-resistant strains [52,53]. However, several studies refute this association because K128T substitution has been found in multiple FLC-susceptible isolates, hence, in this study, we do not consider it as a mutation associated with resistance [25,54]. Nevertheless, it might influence translation efficiency, leading to alterations in protein production as it occurs with nearby mutations such as G129R and Y132F [55].

According to Chow and coworkers, mutations contributing to FLC resistance are clade-specific in *C. auris*, being Y132F and K143R the most predominant in clade I, F126L in clade III, and Y132F in clade IV. However, Y132F in Erg11 is the most prevalent one [26]. As described for isolates of clade IV (South America), we found only one mutation:Y132F. Concerning *C. parapsilosis*, high prevalence of the Y132F substitution was also noted in previous studies [56,57]. In addition, the latter suggested that isolates harbouring this mutation may have a higher propensity to cause clonal transmission and to persist in nosocomial settings [58]. Regarding our isolates, the Y132F substitution was detected in four out of six *C. parapsilosis* isolates. However, considering the dates of collection of the isolates, It does not seem to be a clonal spread (**Table S3**).

Although previous studies indicate that the Mrr1 substitutions I283R, R479K, G583R, A854V, K873N and L986P are associated with FLC and/or VRC resistance, none of our isolates harbored them [59]. Similar to Tac1, residues located near the C terminus (760, 761, 803, 956, and 966) might contribute to azole resistance [47].

This study also illustrates that acquired azole resistance commonly relies on combined molecular mechanisms in clinical isolates. In addition to amino acid substitutions in Erg11, six of the 19 isolates also displayed active efflux activity. Interestingly, in two isolates lacking resistance-related mutations, efflux pumps activity was observed. Considering that efflux pumps play a key role in azole resistance, this might be the explanation for the observed phenotype. However, a further study evaluating the expression of all genes involved in efflux pump activity is required.

Regarding the isolates in which no mechanism for resistance was found, namely CAAL-A-E, CAAU-D, CAAU-E and CAPA-A, six novel Tac1 or Mrr1 substitutions (G52R, V366del, F682T, F683L, T695S and P229A) were identified in three of the five isolates. Moreover, four novel Erg11 substitutions (K22E, Q38T, F72V, Q77S) in the FLC resistant *C. albicans* (CAAL-B, CAAL-C) and *C. auris* (CAAU-B) were found. The putative role of these substitutions remains to be investigated. In isolates CAAL-A-D and CAAU-E, a different mechanism to those evaluated here should confer resistance; further analysis is ongoing.

Our study has some limitations. For instance, the AST data obtained for antifungals other than FLC are not complete and were not performed by BMD. The molecular basis of FLC resistance was investigated only by analysis of the *ERG*11, *TAC*1, and *MRR*1 genes, while other mechanisms conferring resistance, such as sterol composition and gene expression, were not investigated due to lack of funds. Finally, the clinical description of the patients was missing and will be reported alongside other patients infected by *Candida* antifungal resistant strains elsewhere.

Recently, the WHO published a list of priority pathogenic fungi (FPP) which includes *C. albicans, C auris* (Critical group), and *C. parapsilosis* (high group). The WHO describes that to overcome the lack of knowledge on infections caused by these fungi, more data and evidence on fungal infections and antifungal resistance to inform and improve response to FPP is needed [60]. Although our study does not have a large number of isolates and does not include other healthcare institutions, we provide evidence of endemic isolates harboring resistance mutations to one of the most used molecules in the hospital setting.

In conclusion, we describe the presence of *Candida* spp isolates harboring Erg11 FLC resistant-related substitutions (Y132F, K143R and T220L) in patients admitted to a Colombian hospital. Although FLC-resistance rates differ significantly between countries and individual health facilities, the resistance rate in our study is relatively high, emphasizing the need for active surveillance to prevent further expansion of FLC-resistant *Candida* spp isolates in the clinical setting.

## Data Availability

All data produced in the present study are available upon reasonable request to the authors

## Ethics approval and consent to participate

The research and ethics committee of the Hospital Universitario San Ignacio (HUSI) approved this study (no. FM-CIE-8053-14). All patients are anonymized and only the code of isolates was transferred for this investigation. Therefore, no informed consent was required.

## Author contributions

AP carried out the experiments; Y.V-C contributed to the development of efflux pumps protocol; AC-G, and AP analyzed the data; AC-G wrote the main manuscript; AP, SV-B, BA, and CP-G review and editing; and CP-G conceived the experiments and managed the resources. All authors have read and agreed to the published version of the manuscript.

